# Brain activations during execution and observation of visually guided sequential manual movements in autism and in typical development: A study protocol

**DOI:** 10.1101/2023.12.10.23299792

**Authors:** Erik Domellöf, Hanna Hjärtström, Anna-Maria Johansson, Thomas Rudolfsson, Sara Stillesjö, Daniel Säfström

**Affiliations:** Department of Psychology, Umeå University, Umeå, Sweden; Department of Health, Education and Technology, Luleå University of Technology, Luleå, Sweden; Centre for Musculoskeletal Research, Department of Occupational Health Sciences and Psychology, University of Gävle, Gävle, Sweden; Department of Integrative Medical Biology, Umeå University, Umeå, Sweden

## Abstract

Motor issues are frequently observed accompanying core deficits in autism spectrum disorder (ASD). Impaired motor behavior has also been linked to cognitive and social abnormalities, and problems with predictive ability have been suggested to play an important, possibly shared, part across all these domains. Brain imaging of sensory-motor behavior is a promising method for characterizing the neurobiological foundation for this proposed key trait. The present functional magnetic resonance imaging (fMRI) developmental study, involving children/youth with ASD, typically developing (TD) children/youth, and neurotypical adults, will investigate brain activations during execution and observation of a visually guided, goal-directed sequential (two-step) manual task. Neural processing related to both execution and observation of the task, as well as activation patterns during the preparation stage before execution/observation will be investigated. Main regions of interest include frontoparietal and occipitotemporal cortical areas, the human mirror neuron system (MNS), and the cerebellum.

## Introduction

Motor performance difficulties are highly prevalent in children and youth with autism spectrum disorder (ASD), adding to the challenges in daily life functioning [1]. The etiology is not clear, although converging evidence suggests that predictive control deficits may play a part in the presented movement problems [2–4]. Importantly, a similar connection has been made between predictive ability and autism core symptoms such as deficits in social interaction [5–6]. As autism is a neurodevelopmental condition, linked to early impact on the developing brain, it is thus possible that similar neural mechanisms related to predictive behavior operate in an atypical manner across different domains. Still, very little is known about parallel neural activity during behavioral performance requiring predictive control in this population.

Investigating goal-directed motor behavior in young individuals with ASD is a useful way to collect quantitative and reproduceable evidence for impaired predictive functioning. Brain imaging studies of action execution can provide a neurobiological characterization of this potentially defining trait in autism but are very rare. In a seminal study, Mostofsky and colleagues reported that basic motor execution (finger tapping) was linked to atypical brain activations in the cerebellum and the supplementary motor area compared with typically developing (TD) children [7]. This suggests a neurological base for difficulties with motor performance. However, as far as we know, there are no studies to date that have investigated more complex, visually guided goal-directed movements using functional brain imaging in individuals with ASD.

In everyday life, we spend a significant amount of time using our hands in various goal-directed activities [8]. Typically, these activities are comprised of a series of sequentially linked actions, e.g. we reach for and grasp an object in order to do something with it. An impairment in the planning of movement and/or predictive movement control will make the execution of sequential manual tasks less smooth and efficient. Performance of action sequences has been linked to activity in frontoparietal and occipitotemporal circuits in neurotypical individuals [9–10]. Whether individuals with ASD engage these functional brain networks in the same way is currently unknown.

Impaired prediction skills may further influence the understanding of actions performed by others. The mirror neuron system (MNS) of the brain, located in frontoparietal brain regions, is activated both when executing and observing goal-directed action [11]. As such, the role of the MNS and extended networks has been targeted in several brain imaging studies related to observation of human biological action in children and youth with ASD, including gestures [12], hand reaching [13], eating and placing [14], and goal-directed action [15]. Findings from these studies are divergent and suggest either atypical activation in young participants with ASD compared with TD controls or no global differences between ASD and TD controls.

Taken together, more knowledge is needed regarding shared neural processing related to action execution and action observation within the MNS both in general and in individuals with ASD. To properly investigate this, brain imaging studies combining execution and observation of a goal-directed movement involving motor planning and predictive control are required. Brain activity associated with action execution and observation in the MNS is possibly influenced by the ability for predictive control. As outlined above, impaired predictive control has been suggested as a salient phenotype in autism. Using a thorough methodology and a complex task (e.g. a goal-directed sequential action), it is possible that more clear-cut differences in patterns of MNS activation between ASD and TD individuals will be found compared to previous investigations. Finally, given the early onset of impairments in ASD, it is important to involve a developmental perspective in this research endeavor. To this end, the present study aims to investigate brain activations during execution and observation of visually guided sequential manual movements in one group of children/youth with ASD (12-16 years), one age-equivalent group of TD children/youth, and in one group of neurotypical adults (18-40 years). In addition, the children/youth will be asked to participate a second time one year after the first experimental run.

### Study aims

Brain activations will be measured by functional magnetic resonance imaging (fMRI) generating blood oxygen level dependent (BOLD) contrasts for activity in different brain areas during (i) execution of a visually guided, goal-directed sequential (two-step) manual task, and (ii) observation of a video showing a hand executing the sequential manual task. The following main aims will be pursued with regard to differences between children/youth with ASD, TD children/youth, and neurotypical adults:

1. Investigate brain activity during the preparation stage before executing a sequential manual task
2. Investigate brain activity during the preparation stage before observing a sequential manual task
3. Investigate overlapping brain activity during the respective preparation stage before executing and observing a sequential manual task
4. Investigate brain activity during execution of a sequential manual task
5. Investigate brain activity during observation of a sequential manual task
6. Investigate overlapping brain activity during execution and observation of a sequential manual task

Additional aims include exploring outcomes and group differences associated with behavioral execution measures collected in the scanner (latency, duration) in relation to brain activity (including motor variability in ASD), behavioral measures collected outside the scanner (fluid intelligence, working memory, cognitive planning, fine motor performance) in relation to brain activity, cerebral functional connectivity in resting state functional MRI (rs-fMRI) in relation to functional brain activity, and diffusion tensor imaging (DTI) and Synthetic MR (SyMR), respectively, in relation to brain activity. Moreover, employing a longitudinal approach will allow investigation of developmental changes in the children/youth in relation to the stated aims.

#### Hypotheses

Anticipatory planning of movement involves associated activity in frontal brain areas for processing of action plans, and parietal brain areas for monitoring of action outcomes [16]. Planning of object-directed action sequences has been found to further engage occipitotemporal cortex [9–10]. In relation to the main aims, we thus expect to find preparative processing in frontoparietal and occipitotemporal cortical areas, with possible overlap between task execution and observation. The MNS, involving premotor cortex, inferior parietal lobe, primary motor and somatosensory cortex [17], is expected to be engaged during task observation. We further hypothesize that while regional activations may be largely similar between groups, intensity and direction of activity will differ depending on ASD diagnosis (ASD vs. TD) and age (TD vs. adults). In addition, both structural cerebellar abnormalities and atypical motor-related functional activation of the cerebellum are commonly reported in ASD [7]. Action prediction depends on the integrity of a cortico-cerebellar loop, from posterior parietal cortex via the cerebellum and back to premotor and motor brain regions [4, 18]. The cerebellum has further been implicated in functions outside the sensorimotor domain, including executive functioning and social cognition [19]. Therefore, we further expect different cerebellar activation patterns in children/youth with ASD compared to TD controls.

## Materials and Methods

### Study design

The study involves both a cross-sectional and longitudinal approach to investigate differences and commonalities in neural processing during execution and observation of visually guided goal-directed sequential manual movements at two time points (T1-T2) in children/youth with ASD and TD children/youth, also as compared with neurotypical adults (gold standard for fully developed task-related predictive ability) at T1. Presently, participant recruitment and data collection have been initiated, but as requested for a study protocol and in compliance with PLOS data policy, no data has been included. Table 1 outlines the assessments covered by the study, and the timeline for the study (Gantt chart) is depicted in Figure 1.

**Figure 1.**
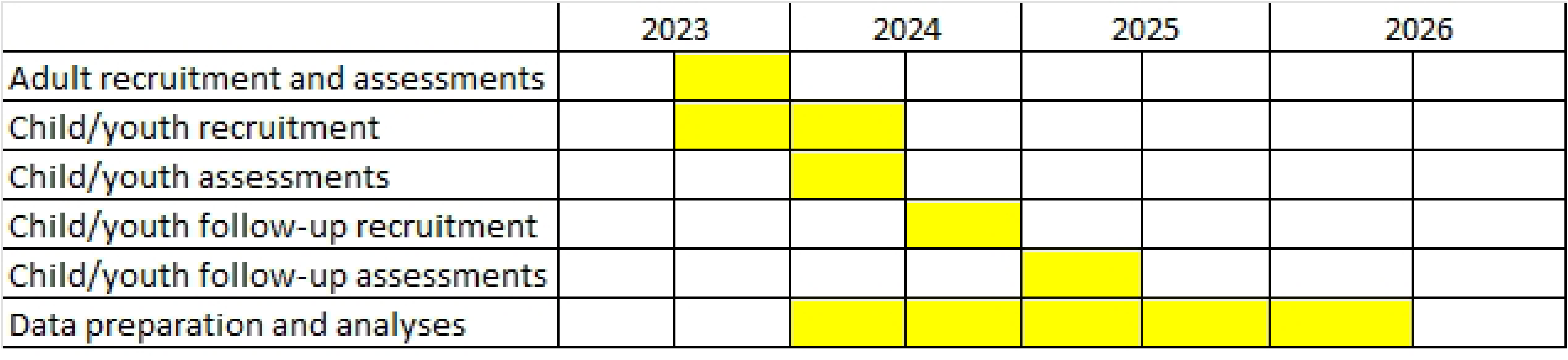
Timeline for study recruitment, assessments, and data preparation and analyses.

**Table 1.** Cognitive, motor and brain imaging assessments. T1, assessment time point 1; T2, assessment time point 2.

### Ethical approval

The study was approved by the Swedish Ethical Review Authority (ref: 2022-01513-01) and is conducted in accordance with the Declaration of Helsinki.

### Participants

A total of 30 neurotypical adults (18-40 years) and 40 children/youth (20 ASD, 20 TD; 12-16 years) will be recruited. The 12–16 years age span was chosen because these children/youth are old enough to manage a complex sequential motor task in the scanner but have not yet developed an adult level of dexterity. Sample sizes are in keeping with, or larger than most similar previous fMRI studies and can be considered acceptable in terms of generating sufficient statistical power [20]. Inclusion criteria for the adults and the TD children/youth are language fluency in Swedish, normal or corrected-to-normal vision, and no diagnosed neuropsychiatric or motor disability. Inclusion criteria for the children with ASD are an ASD diagnosis established within the Swedish health care system, no diagnosed intellectual disability, and language fluency in Swedish. Exclusion criteria for all groups are non-compliance with the testing protocol, metal objects in the body (implanted or otherwise), or pregnancy. A specific exclusion criterium for the adults, but not the children/youth, is left-handedness. The children with ASD are recruited through records from the Habilitation Centre, Region Västerbotten, Sweden. Families are contacted by the Habilitation Centre and asked if the researchers may contact them and inform them about the study. If they agree, the families receive a participant information letter followed by a telephone call to ensure verbal information and answers to questions. The TD children and adults are recruited through advertisements in the Umeå region and by convenience sampling. Families of TD children and adult participants also receive a participant information letter followed by an e-mail or a telephone call. Of particular relevance for children/youth with ASD, the information letter also refers to a dedicated home page with pictures and videos of study premises, people involved in the study, and what participants are supposed to do. Recruitment started October 2023. Assent from the child and written, informed consent from both parents/caregivers are collected for all child participants. Written, informed consent is also collected from children/youth aged 15 years or older and from adult participants.

### Equipment

For the fMRI execution task, two identical sets of custom-made wooden equipment were built (Figure 2), one for practicing and one for use in the MR scanner. The equipment, purposely designed for the study, consists of a wooden box with two compartments where a standard fMRI 4-button response box (Current Designs) can be fitted. The top covering the wooden compartment has two vertically aligned holes to the left side (for a right-handed participant), corresponding to buttons 1 and 4 on the fMRI response box underneath, and a T-shaped frame made of braided wool yarn glued to the surface in a 30⁰ clockwise rotational angle to the right side. Adjustment to a left-handed participant can be made by exchanging the top for another where the holes are situated to the right, the T-frame to the left, and using the second compartment of the wooden box. The T-frame corresponds to a wooden object consisting of a cube (1.5 x 1.5cm) with four cylinders (length: 4cm, diameter: 1.5 cm) glued to the cube (Figure 2). The cylinders are in four different colors: One yellow (corresponding to the vertical part of the “T”), one black (horizontal right side of the “T”, facing upwards toward the top button), one red (horizontal left side of the “T”, facing downwards toward the button below), and one blue (facing the participant).

**Figure 2.**
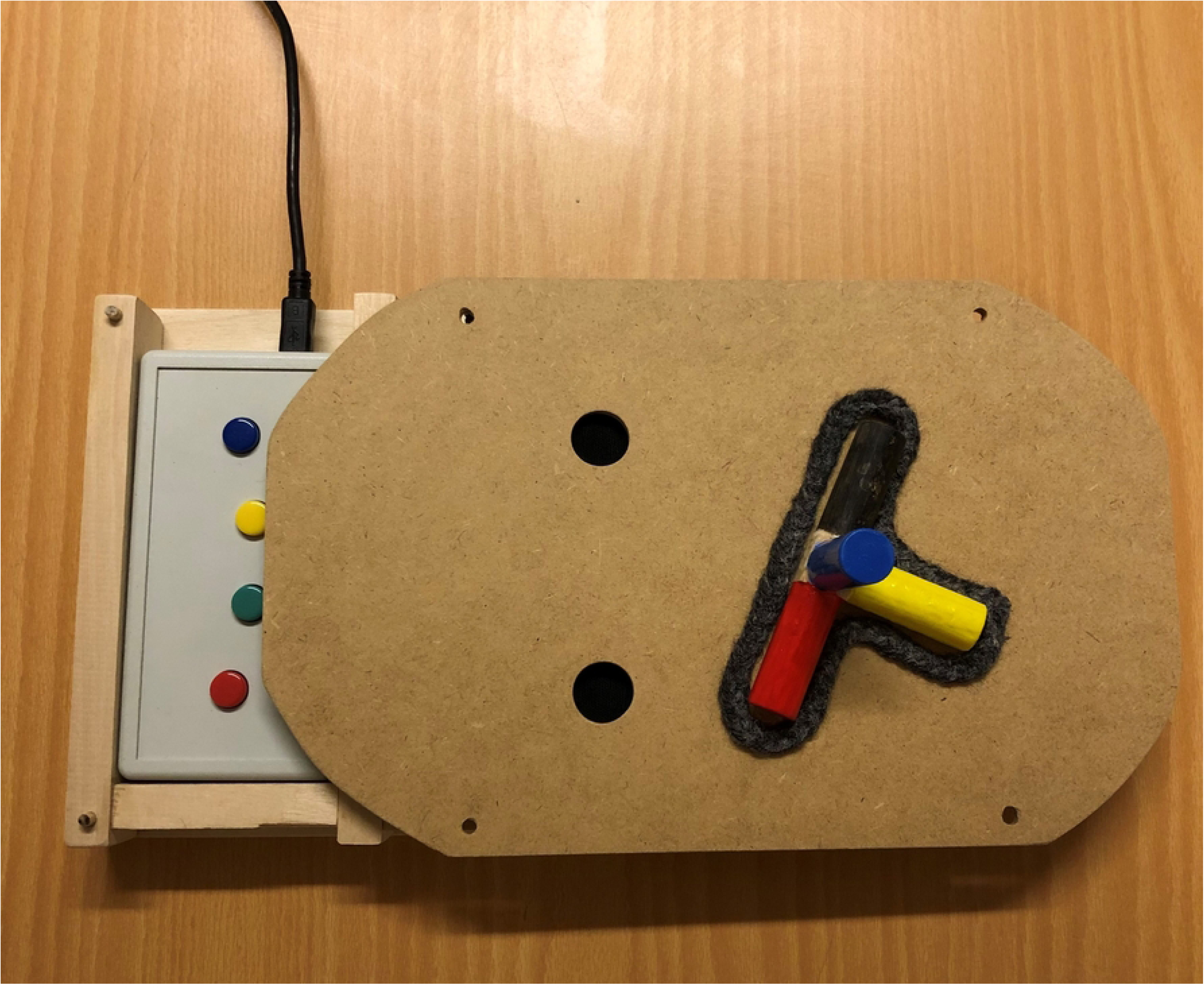
The equipment (for right-handed participants): wooden box with fMRI 4-button response box fitted into compartment, top display with holes corresponding to buttons 1 (blue) and 4 (red) and T-shaped frame with the wooden object inserted.

### Experimental task

The fMRI execution task consists of, holding the yellow cylinder, using the cylinders of the wooden object to sequentially press the top button first, then the second button below, and then return the object to the frame. The top button is always pressed by the black cylinder, while the second button can be required to be pressed by either the black, red, blue or yellow cylinder. Pressing black-black, where no rotation of the object is needed between buttons, requires less motor planning and constitutes the baseline (“baseline trials”). Pressing black-red/blue/yellow requires a complex rotation of the object, and therefore increased motor planning is demanded for correct performance (“complex trials”).

For the fMRI observation task, short video clips of a hand, holding the yellow cylinder, moving the wooden object to press black-black, black-red, black-blue, and black-yellow, respectively, were recorded. The videos show the execution of the trials from the same view as when the participants perform the task themselves (first-person view). To stimulate engagement in the observation task, two exclusive videos for each of the four respective trial variants were recorded (i.e. offering a slight variation between videos). Schematic illustrations including measures of the equipment (top display and compartment) and the wooden object are added in Supplementary Information (Figures S1-S3), along with one example of the observation task videos (S4).

As shown in Figure 3, the task structure is as follows: First, delivered by a dedicated custom-made PsychoPy script, the participant receives a cue depicting (i) condition (execution/hand or observation/eye) and (ii) a schematic image of the top display (the object with the differently colored cylinders in starting position and the two holes) with the color of the holes indicating which of the colored cylinders that are required to press the buttons (e.g. black-blue). During cue presentation (5s), which constitutes the preparation stage, the participant is asked to either think about how to perform the movement (execution task) or prepare to watch a video of the movement (observation task). The cue is followed by a delay (2-10s inter-stimulus-interval, ISI, including a probe to either look at hand or continue to watch the screen), a subsequent auditive start signal, and then either execution or observation of the manual task.

**Figure 3.**
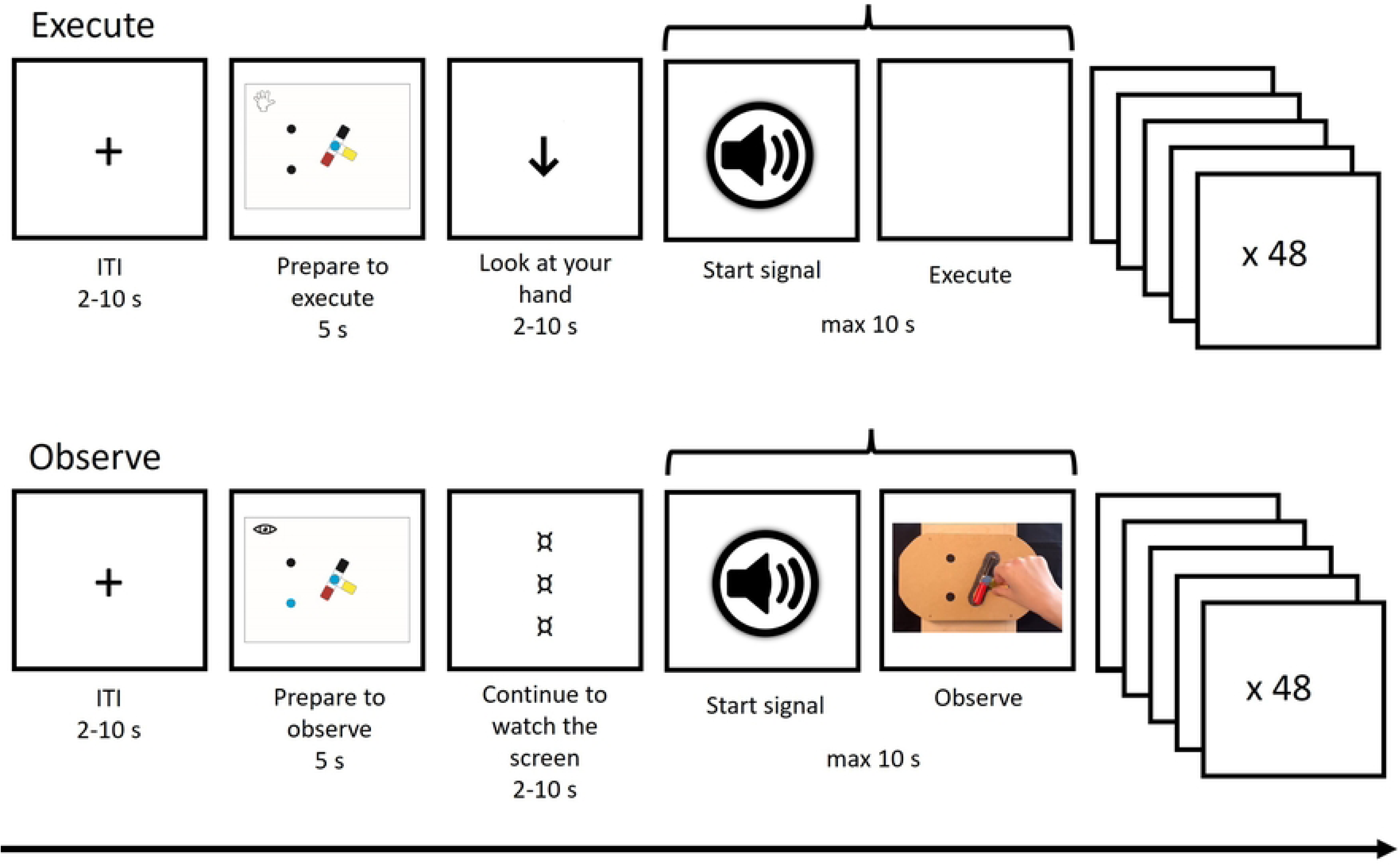
The fMRI task structure and trial procedure. ITI, inter-trial interval.

In the execution condition, responses (button presses) are logged by the fMRI response box underneath the top display, with the final response (second hole) determining end of trial. If no response is logged within 10s, the trial ends automatically. After this, another delay (2-10s inter-trial-interval, ITI) is administered before the next cue is presented. A total of 48 trials, 24 baseline and 24 complex, are administered for execution and observation, respectively.

### Data collection

Participation in this study comprises two sessions (Figure 4), one practicing session (1.5 hours in total) and one fMRI session (2 hours in total). Time between practicing and fMRI may vary but is no longer than 2 weeks. Before data collection, the participants (and/or caregivers of young participants) complete a brief questionnaire providing demographic information and a modified version of the Edinburgh Handedness Inventory [21]. To investigate the presence of autistic traits among the adults, adult participants are asked to fill out a screening questionnaire (Autism Spectrum Quotient, AQ).

**Figure 4.**
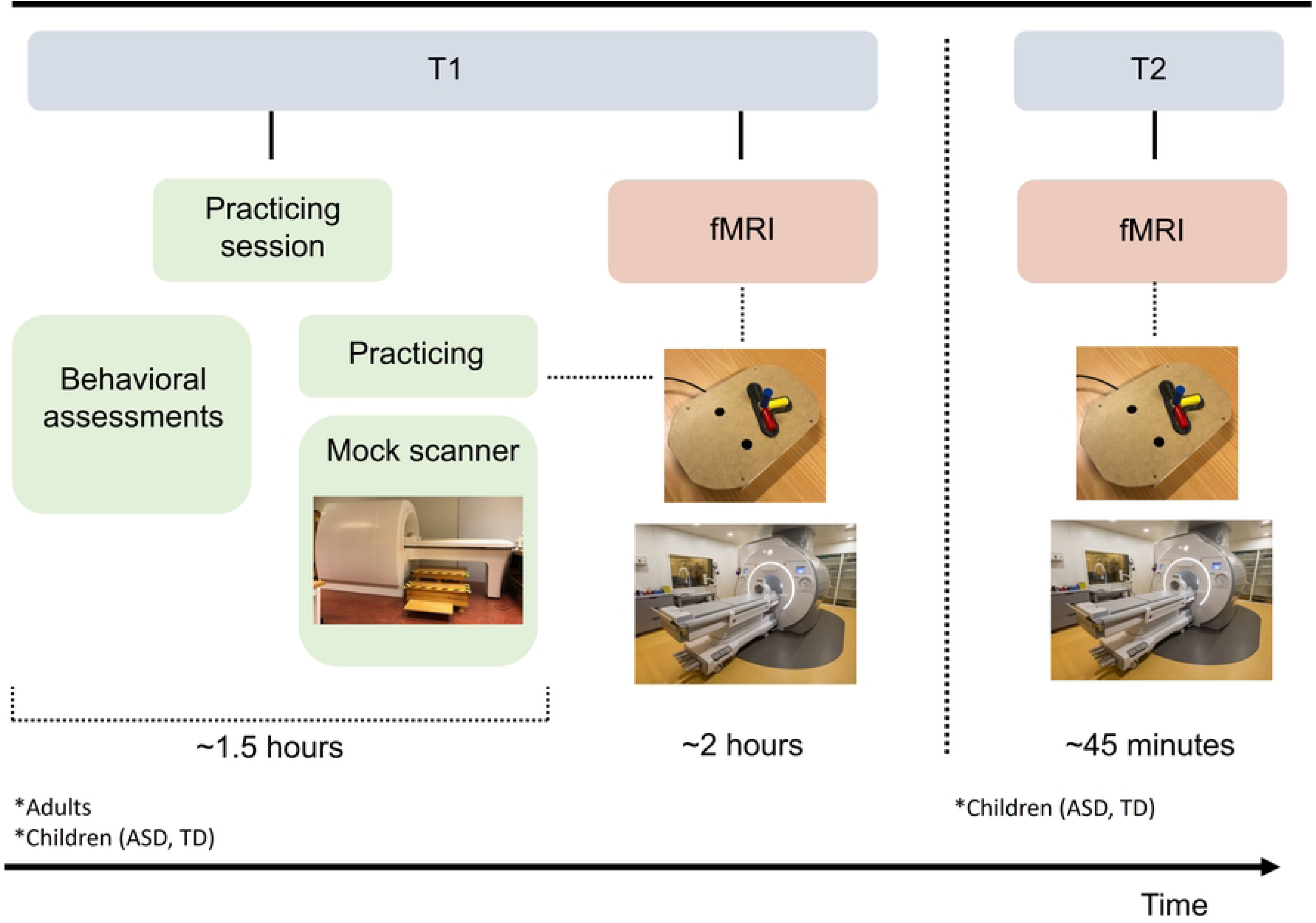
Overview of the study sessions. * denotes populations that are included at assessment time point 1 (T1) and 2 (T2), respectively. Photo of MR scanner by Mikael Stiernstedt.

#### Practicing

The practicing session takes place at the Department of Psychology, Umeå University, and include (i) priming in a mock scanner (realistic model of a real MR scanner with recorded noise), (ii) learning the sequential manual task and the fMRI protocol, and (iii) behavioral assessments (Table 1). The priming in the mock scanner allows the participants to familiarize themselves with lying in a MR scanner. Also, if the participant should experience claustrophobia, participation can be ruled out from the very beginning. For practicing the sequential manual task, a wooden equipment (see Equipment above) and a 4-button response box (same as the fMRI response box but adapted for use outside the scanner) attached to a laptop computer are utilized. The same PsychoPy script as described above is employed, although total number of trials is reduced to 8 trials, 2 baseline and 6 complex (2 of each color red/blue/yellow), for execution, and 4 trials, 1 baseline and 3 complex (1 of each color red/blue/yellow) for observation. Prior to practicing, the participant receives verbal instructions (supported by pictures of the cues). If needed, additional instructions and support are also provided during the practicing. In addition, the participant is given an illustrated instruction sheet to take home and read before the fMRI session. For execution trials, particular emphasis is placed on thinking about how to perform the movement during the execution/hand cue, on looking down at the top display/own hand during the ISI, and on waiting for the auditive signal to start. These instructions are important to avoid simply using color pattern memorizing and to avoid contaminating task execution brain engagement with activity due to re-directing gaze from the screen to the hand. For observation trials, particular emphasis is put on holding the hand still while watching the video clip, to avoid brain activity linked to hand use contaminating task observation activations. Finally, behavioral assessments (Table 1) are carried out in a dedicated test room by, or under supervision by, a licensed psychologist (ED). Both cognitive and fine motor general functioning are important for the manual task studied. Associations between cognitive functions and motor planning and performance have also been reported in ASD [22–23]. Therefore, these assessment outcomes are collected for comparative purposes and for use as covariates in the analyses.

#### fMRI investigation

The fMRI session takes place at the Umeå center for Functional Brain Imaging (UFBI)/MR-center at Umeå University Hospital, utilizing a 3T MR scanner. Prior to entering the scanner, the instructions are once more gone through, and any minor visual impairment is corrected by fitting of dedicated fMRI goggles. The MR staff additionally inform the participant about the MR procedure. To ensure protection from the loud scanner noise and for audio purposes, the participant is provided with earplugs and ear protectors. The participant is placed in the MR scanner with the head raised 27°, which enables visual guidance of the hand during execution trials and visibility of a computer screen behind the scanner bore (screen size 88.2 x 48.5cm). The computer screen, which is visible through a tilted mirror affixed to the head coil (Figure 5), is used to give the instructions during the experiment and to show videos of the task during observation trials.

**Figure 5.**
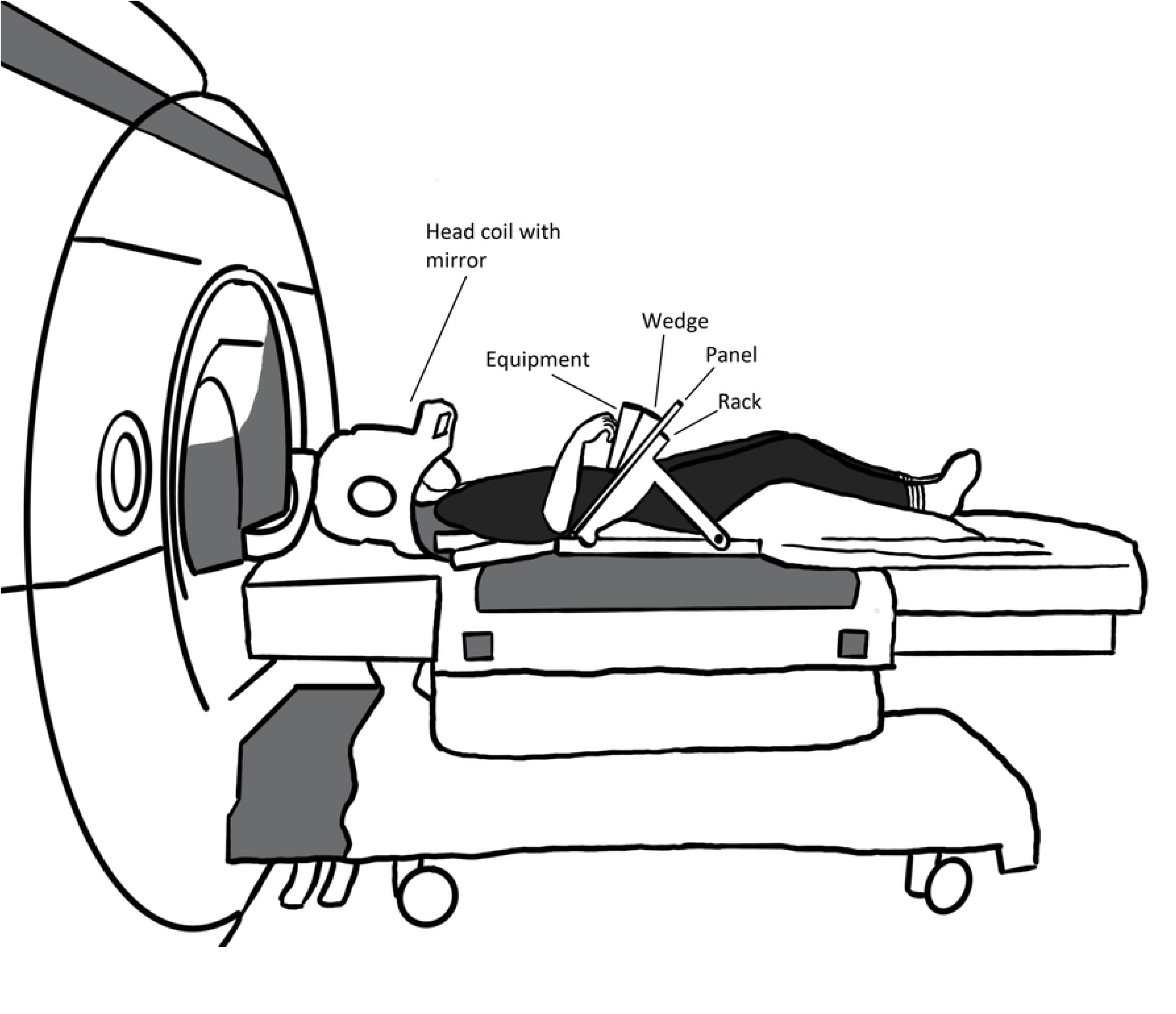
Illustration of a participant lying on the MR bunk with head coil, rack, wedge and equipment fitted, ready to go into the scanner.

A custom-made wooden rack with a screwed on (using brass screws) arched wooden panel, is fitted on top of the participant. A 15° wooden wedge holding the equipment (secured by velcro) is fitted onto the panel, allowing individual adjustment of the equipment in either direction. All fittings are made so that execution of the task can be performed comfortably by each participant. All participants first perform T1-weighted structural imaging (4:11 min) and fMRI imaging (3×14 minutes), followed by T2*-weighted imaging (2:05 min), rs-fMRI (6 min), DTI (5:20 min), and SyMR (5:51 min). During scanning, participant heart rate and respiration are monitored, and the participant can at any time communicate with the MR staff by using an emergency squeeze ball. Instructions are given to squeeze this ball primarily if feeling discomfort or needing to stop the scan for any reason.

### MRI data acquisition and preprocessing

Image acquisition is made using a 3T GE Discovery MR 750 scanner (General Electrics). T2-* weighted images are obtained using a single shot gradient echo planar imaging (EPI) sequence used for Blood-Oxygen-Level-Dependent (BOLD) imaging. The following parameters are used for the imaging sequence: echo time: 30ms, repetition time: 2000ms (37 slices acquired), flip angle: 80°, field of view: 25×25cm, 96×96 matrix and 3.4mm slice thickness. A TDI AIR 48 channel SIGNA Premier XT head coil is used. Ten dummy scans are collected and discarded before the start of the fMRI data collection to allow for equilibrium of the BOLD-signal.

T1-weighted images are obtained using a 3D fast spoiled gradient echo sequence (MP-rage) in axial orientation. The parameters are: echo time: 30ms, flip angle: 8°, field of view: 24 x 24cm and 0.5mm slice thickness.

Resting-state fMRI data is acquired using the following parameters: echo time: 30ms, repetition time: 2000ms (37 slices acquired), flip angle: 80°, field of view: 25×25cm, 96×96 matrix, slice thickness: 3.4mm, slice spacing: 0.5mm.

A 2D MaGiC sequence is used to collect synthetic MRI. The parameters are: Number of echoes: 2, repetition time: 4803ms (31 slices acquired), field of view: 24 x 24cm, slice thickness: 4mm, slice spacing: 1mm.

All images will be corrected for slice timing and realigned to the first image volume of the series. All images will be spatially smoothed through DARTEL [24]. The individual T1-images will be segmented, and a group-specific mean template and individual flow fields will be created using the DARTEL algorithm. All T1-images will then be normalized using the group-specific template and adjusted to the anatomical space defined by the MNI atlas using SPM12 (Wellcome Department of Cognitive Neurology, UK). All images will be smoothed using 8.0mm full width and half maximum (FWHM) Gaussian Filter Kernel. All data will be high-pass filtered (128s cutoff period) to remove low frequency noise. The fMRI data will be analyzed using SPM12 implemented in Matlab (R2014b) (Mathworks, Inc., USA) and run through an in-house program (DataZ).

### Outcome measures

#### Brain imaging

The fMRI-experiments will generate blood oxygen level dependent (BOLD) contrasts for task-related activity in different brain areas during execution, observation and the preparation stage (before execution/observation). The T1-weighted structural images will be used to validate a correct anatomical localization of significant clusters of activation and local maxima. To elucidate normal and abnormal functional connectivity between different brain regions, we will use resting-state fMRI (rs-fMRI) that measure spontaneous low-frequency fluctuations in the BOLD signal, where functionally related regions may exhibit correlations in activity. The structural connectivity between brain regions will be investigated by diffusion tensor imaging (DTI) that measures the anisotropic diffusion of water molecules in white matter fibre tracts (tractography). Finally, synthetic MR (SyMR) is included in the study protocol because it rapidly measures tissue properties that enables several different contrasts to be generated. These contrasts will be used to investigate structural brain differences between groups, and developmental changes enabled by our longitudinal approach.

#### Behavior

During the practicing session we will quantitatively measure both cognitive and fine motor general functioning. The cognitive measurements include fluid intelligence (Wechsler Adult Intelligence Scale, Fourth Edition/WAIS-IV and Wechsler Intelligence Scale for Children, Fifth Edition/WISC-V), working memory (WAIS-IV, WISC-V and Corsi Block-Tapping Test) and cognitive planning (Delis-Kaplan Executive Functions System/D-KEFS). Fine motor performance level will be measured with the Grooved Pegboard Test. During the fMRI session we will measure behavioral parameters during task execution, such as the latency between the start signal and the first button press, and the duration between button presses.

### MRI data analysis

General linear models will be created based on eight effects of interest (*preparation stage:* execute baseline, execute complex, observe baseline, observe complex; *execution/observation:* execute baseline, execute complex, observe baseline, observe complex) and effects of no interest related to events for ITI and ISI. All regressors except the six movement parameters will be convolved with a hemodynamic response function. Each event will be modeled from onset to offset. The time when an event appears on the screen is defined as onset, and response times will be used as offset. The durations will be variable to allow the analyses to capture the complete process of preparation, execution and observation, which could differ from time to time.

In the first level, estimations will be made separately for each participant, and a *t*-contrast will be created for all effects of interest. The *t*-contrasts defined at the first level will then be taken into a second-order group level analysis, and inferential statistics (*t*-tests, conjunction analyses, ANOVAs and exploratory whole-brain correlations) will be used to answer the research questions.

Additionally, measures from the behavioral assessment, age, sex and response times will be used as covariates in the analyses to evaluate effects related to individual differences. Preprocessed resting state data and DTI data will also be analyzed related to the research questions. Additionally, exploratory analyses will be made using synthetic MR data. Finally, the in-house program DataZ (run through SPM 12) will be used to analyze longitudinal data associated with the main task, DTI, rs-fMRI and SyMR.

The statistical analyses will be corrected for multiple comparisons (*p* < 0.05 FDR, *p* < 0.05 FWE) at the voxel level.

### Behavioral statistical analyses

All behavioral measures (e.g. fine motor performance level or latency between the start signal and the first button press) will be analyzed using ANOVAs for comparisons of between-subject factors (our three participant groups) and within-subject factors (such as baseline trials vs complex trials). In all tests, p < 0.05 will be considered statistically significant.

### Data management

The study involves information classification, risk– and vulnerability analysis based on best practice according to the Swedish Civil Contingencies Agency, and a plan for data storage/liquidation. In keeping with the European Union General Data Protection Regulation (GDPR) and national guidelines for the storing of personal data, an exclusive secure file area for the study has been set up, governed by the Umeå University IT department. Data collection has been started but no datasets have yet been generated or analyzed. In accordance with GDPR and Data Protection Act 2018, data will be de-identified as soon as practically possible. All data are pseudonymized, using a specific study code for each participant. Data obtained from the MR scanner sessions are stored in a secure computer database kept at the UFBI. Additional background and behavioral data collected outside the scanner are securely kept in a safety cabinet in a locked laboratory at the Department of Psychology, Umeå University. Only those directly involved in the project have access to data. De-identified data originating from this study will be made available upon completion and shared with the scientific community.

## Discussion

Motor performance difficulties are frequently observed in ASD and have also been linked to autism core symptoms such as deficits in social interaction. Possibly, an impaired predictive ability is the key common denominator across these different domains. To investigate this, sensory-motor behaviors are a particularly useful source of information because they are possible to accurately quantify and reproduce. Despite this usefulness, and despite the immediate importance of understanding the core neurodevelopmental processes in autism, there is to date a shortage of fMRI studies exploring actual movement performance in individuals with ASD. In particular, there is a lack of studies involving direct eye-hand coordination, more complex movements that require predictive ability, and a developmental (longitudinal) design. One obvious explanation for this sparsity is the challenges involved in investigating movement using a method where participants, preferably lying supine in the scanner, should move as little as possible.

There are, however, an increasing number of fMRI studies proving the feasibility of exploring actual goal-directed eye-hand action in adult participants [9, 25]. Through these types of studies, we can gain important knowledge about how brain areas supporting visual processing, motor planning, sensory-motor integration and predictive control are engaged in actions that mimic those that we perform in everyday life. At present, there is also a growing body of evidence showing that children and adolescents, both TD and those with neurodevelopmental conditions, successfully can partake in fMRI studies including those involving a motor task [7, 26]. In the framework of the present study, learning more about the neurobiological characterization of goal-directed sensory-motor behavior, predictive functioning and the observation of other’s action in young participants with ASD, preferably over time, could add importantly to the understanding of core issues in autism.

## Authors’ contributions

<u>Conceptualization</u>: Erik Domellöf, Hanna Hjärtström, Anna-Maria Johansson, Thomas Rudolfsson, Sara Stillesjö, Daniel Säfström

<u>Data curation</u>: Erik Domellöf, Hanna Hjärtström, Anna-Maria Johansson, Thomas Rudolfsson, Sara Stillesjö, Daniel Säfström

<u>Formal analysis</u>: Hanna Hjärtström, Sara Stillesjö

<u>Funding acquisition</u>: Erik Domellöf

<u>Investigation</u>: Erik Domellöf, Hanna Hjärtström, Anna-Maria Johansson, Thomas Rudolfsson, Sara Stillesjö, Daniel Säfström

<u>Methodology</u>: Erik Domellöf, Hanna Hjärtström, Anna-Maria Johansson, Thomas Rudolfsson, Sara Stillesjö, Daniel Säfström

<u>Project administration</u>: Erik Domellöf, Hanna Hjärtström, Anna-Maria Johansson, Thomas Rudolfsson, Sara Stillesjö, Daniel Säfström

<u>Software</u>: Hanna Hjärtström, Sara Stillesjö

<u>Supervision</u>: Erik Domellöf, Anna-Maria Johansson, Thomas Rudolfsson, Sara Stillesjö, Daniel Säfström

<u>Writing – original draft</u>: Erik Domellöf

<u>Writing – review and editing</u>: Erik Domellöf, Hanna Hjärtström, Anna-Maria Johansson, Thomas Rudolfsson, Sara Stillesjö, Daniel Säfström

## Acknowledgements

We are grateful to the Habilitation Centre, Region Västerbotten, to UFBI, and to the Knut and Alice Wallenberg Foundation for supporting this study. We further thank the MR staff at the Umeå University Hospital and Andreas Johansson for their excellent work regarding the ongoing data collection.

## Supporting Information

**S1 Figure.** Schematic illustration of the equipment top display (1:1 scale).

**S2 Figure.** Schematic illustration of the equipment box with compartments (1:2 scale), and 3D drawing including measures of box base plate and side heights (not to scale).

**S3 Figure.** Schematic illustration of the wooden object (not to scale).

**S4 Video.** Example of observation task video (baseline trial).

## Data availability

Results from this study will primarily be communicated in the shape of international scientific publications (open access), conference contributions, and presentations to any interested audience. We further acknowledge a willingness to share the collected and analyzed data, and a de-identified data file will be open for the scientific community. Individual contrast files for the fMRI data will also be made available upon request.

